# Transmission Dynamics of COVID-19 in Malaysia Prior to the Movement Control Order

**DOI:** 10.1101/2020.02.07.20021188

**Authors:** J Labadin, B.H Hong

## Abstract

This paper focuses on the formulation of a deterministic COVID-19 transmission model by considering the exposed and recovered populations with immunity. The scenario of the simulation is depicted based on the patient zero in Malaysia. The transmission model is found to be able to predict the next confirmed case given a single case is introduced in a fully susceptible population. The mathematical model is developed based on the SEIR model which has susceptible, exposed, infectious and recovered populations. The system of equations which were obtained were solved numerically and the simulation results were analyzed. The analysis includes the impact of the disease if no control is taken.

## Introduction

On 31 December 2019, China notified outbreak pneumonia of unknown etiology in Wuhan City to the World Health Organization (WHO) and it had been identified as a novel coronavirus (SARS-CoV-2) causing the COVID-19. Until 13 January, the first COVID-19 cases that was detected outside of China, was from Thailand [1]. During that early period, there were only 41 cases reported in Wuhan city. The Imperial College London published a report to estimate the potential total number of COVID-19 cases in Wuhan City [2]. In the report, 4000 cases are estimated to have an onset of symptoms by 18 January 2020. The estimation is based on the airport catchment population of Wuhan International airport, which is the volume of international traveler per day. Moreover, a group of researchers has estimated the unreported cases in China from 1 January 2020 to 15 January 2020. The estimated unreported cases are 469 cases by using the Maximum Likelihood Estimation (MLE) [3].

On 20 January 2020, the China’s National Health Commission confirmed that the COVID-19 is a human-to-human infectious disease [4] and ten days later, WHO declared that the disease is of Public Health Emergency of International Concern (PHEIC). Though, PHEIC was declared by WHO, there was yet any banning imposed on travels to and from China at that time. The official name for the disease, initially referred as 2019-nCoV or Wuhan Virus was then announced as COVID-19 on 11 February 2020.

The COVID-19 spreads worldwide by the end of February, of which many countries such as North Korea, Italy, Germany and the UK are facing the outbreak as the reported new cases increased exponentially daily. A month later, WHO declared that the COVID-19 outbreak as pandemic, 71 days after China outbreak. By the 24 March 2020, a total of 398,132 confirmed cases and with 17,330 death cases were reported. More than 190 countries were affected by COVID-19. The lock-down control was declared by many countries to break the transmission chain and control the disease from further spread.

As for Malaysia, the Ministry of Health (MoH) reported the first four confirmed cases of whom are of China nationality on 25 January 2020. These four infected patients were reported to have entered Malaysia from Singapore of which two days earlier, a confirmed case was notified by the Singapore authorities. The first infected Malaysian was reported on 4 February 2020. Based on the patient mobility report before the notification, it was noted that he was in Singapore for a week starting on January 16, 2020, to attend an international conference where some of the conference delegates were reported to have come from China. It was noted that this patient’s symptoms appear on 23 January and he only seeks treatment on January 29 and was referred to the Sungai Buloh Hospital on 2 February 2020.

Malaysia decided on travel restrictions for China visitors from Wuhan and Hubei province on 27 January and this restriction was expanded on 9 February which included two more provinces for China which are Zhejiang and Jiangsu [5]. The first wave of COVID-19 was reported to a total of 22 confirmed cases and most of the cases are imported cases. After 11 days of silence, the 23rd case was reported and then by early March the figure increased seventy-fold to 1624 cases. This massive increment was due to two transmission clusters identified where 57% of them are from the Sri Petaling Tabligh Gathering.

The current understanding of the COVID-19 is that there is a possibility that the novel coronavirus can be asymptomatic. Thus, this paper aims to add insights into the epidemiology of COVID-19 by simulating the transmission dynamics of COVID-19 in Malaysia when one local human was infected.

## Model Formulation

The transmission model was formulated based on the epidemiology aspects of the disease:

a. In this study, Malaysia’s population is divided into the SEIR compartmental model which consists of four compartments: Susceptible (*S*), Exposed (*E*), Infected (*I*) and Recovered (*R*). The total population of Malaysia is denoted as *N* and it is assumed that the population is homogeneous. It is assumed that initially the total population is susceptible, hence *S*_0_ = *N* and *S+E+I+R = N*.
b. Based on the growing number of cases worldwide within a short period, the disease is assumed to spread sufficiently fast that the birth and the natural death rates can be considered as negligible in the model.
c. It is assumed that the COVID-19 is able to transmit between human to human.

The compartmental model is thus depicted in Figure 1. Since the patient zero may be infectious and was mobile and freely interacting then susceptible human becomes exposed with the disease at a transmission rate *β*. Once exposed, the incubation period starts and then after the period ends then those exposed moves to the infected compartment at a rate *ϕ* and when they recovered from the disease then they will move to the Recovered compartment at a rate *γ*. However, there are some infected cases who may die due to the disease with the given rate of *ϵ*.

**Figure 1.**
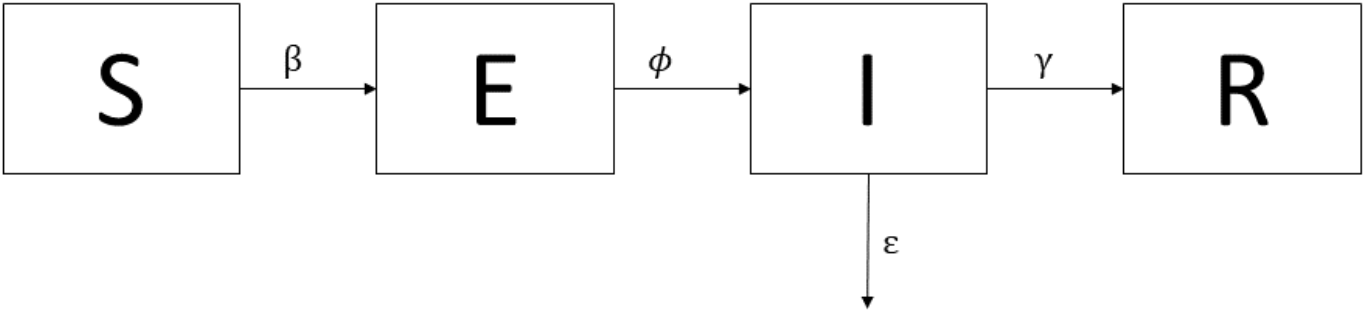
**SEIR Diagram**

The differential equations (equations (1) – (4)) which describe the dynamics of the COVID-19 in human populations are formulated based on the compartmental diagram described earlier in Figure 1.

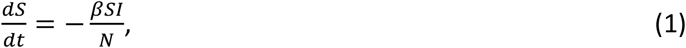

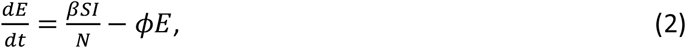

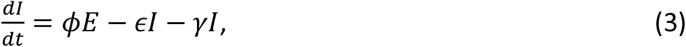

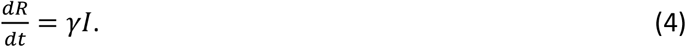

The descriptions of the parameters and the values used for the simulation is shown in Table 1.

**Table 1.** Description of the parameters and their corresponding values used in the simulation

| Parameter | Description | Value | Source |
| --- | --- | --- | --- |
| $N$ | Total human population | 32600000 | [6] |
| $\frac{1}{\phi}$ | Incubation Period | 6.5 | [7] |
| $\beta$ | Transmission rate of $I$ to $S$ | 1.07 | [8] |
| $\frac{1}{\gamma}$ | Infectious Period | 3.6 | [8] |
| $\epsilon$ | Death rate due to COVID-19 | 0.03 | Estimated |

## Model Simulation

The incubation period used for the simulation was 6.5 days which was analyzed by using different distribution approach [7]. The study collected 88 cases that were detected outside Wuhan City, using the on-set date and traveler history to estimate the incubation period of COVID-19. The transmission rate used was 1.07 at the initial stage of the transmission and the infectious period was assigned as 3.6 days [8].

The first confirmed infectious Malaysian case was onset on January 23, 2020 when he came back from Singapore [9]. Hence the initial time (*t*_0_) is set to January 23, 2020 and *I(0)* is 1. *E(0)* and *R(0)* are assigned to 0. The *Rstudio* [10] is used in this study to simulate the SEIR model formulated using the parameter values listed in Table 1.

Figure 2 depicts the simulation of the SEIR model for the first ten days since the on-set date. Based on the result, the number of cases will increase to two persons which indicate that the second case will be confirmed on *t* = 8.74 which is approximately on the 9^th^ day after the on-set date of the first case, that is on the 1^st^ of February 2020 as confirmed in the published report.

**Figure 2.**
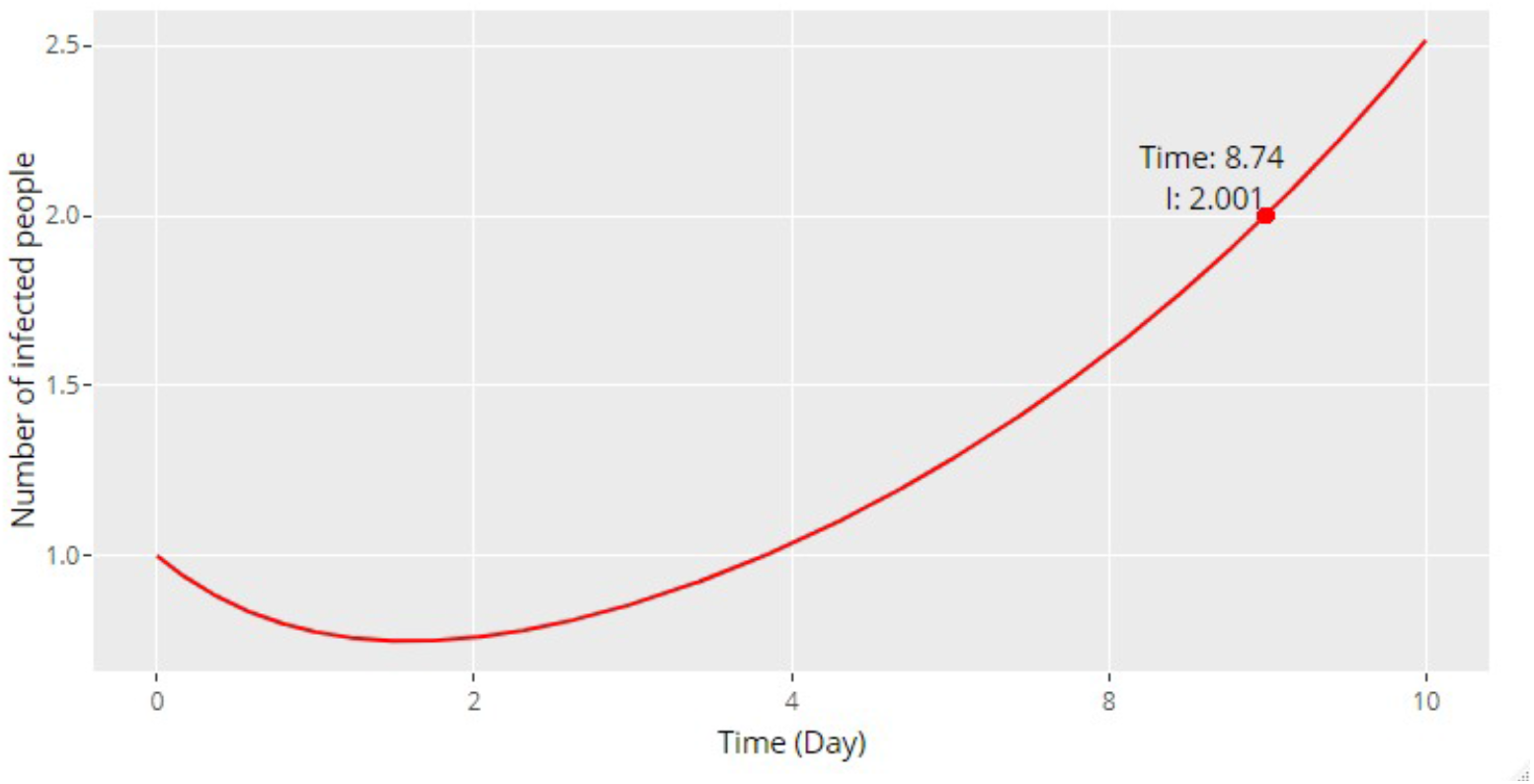
**Simulation of SEIR model in 10 days**

The first wave of COVID-19 outbreak in Malaysia reported 22 cases from 25^th^ January to 17^th^ February and no new case is reported in the next 11 days. The transmission rate, *β* is calibrated and the value is found to be 1.1498, indicating the new transmission rate, *β* used to fit the first wave of the outbreak. Figure 3 illustrated the model fit and the cumulative confirmed cases 25^th^ January to 17^th^ February.

**Figure 3:**
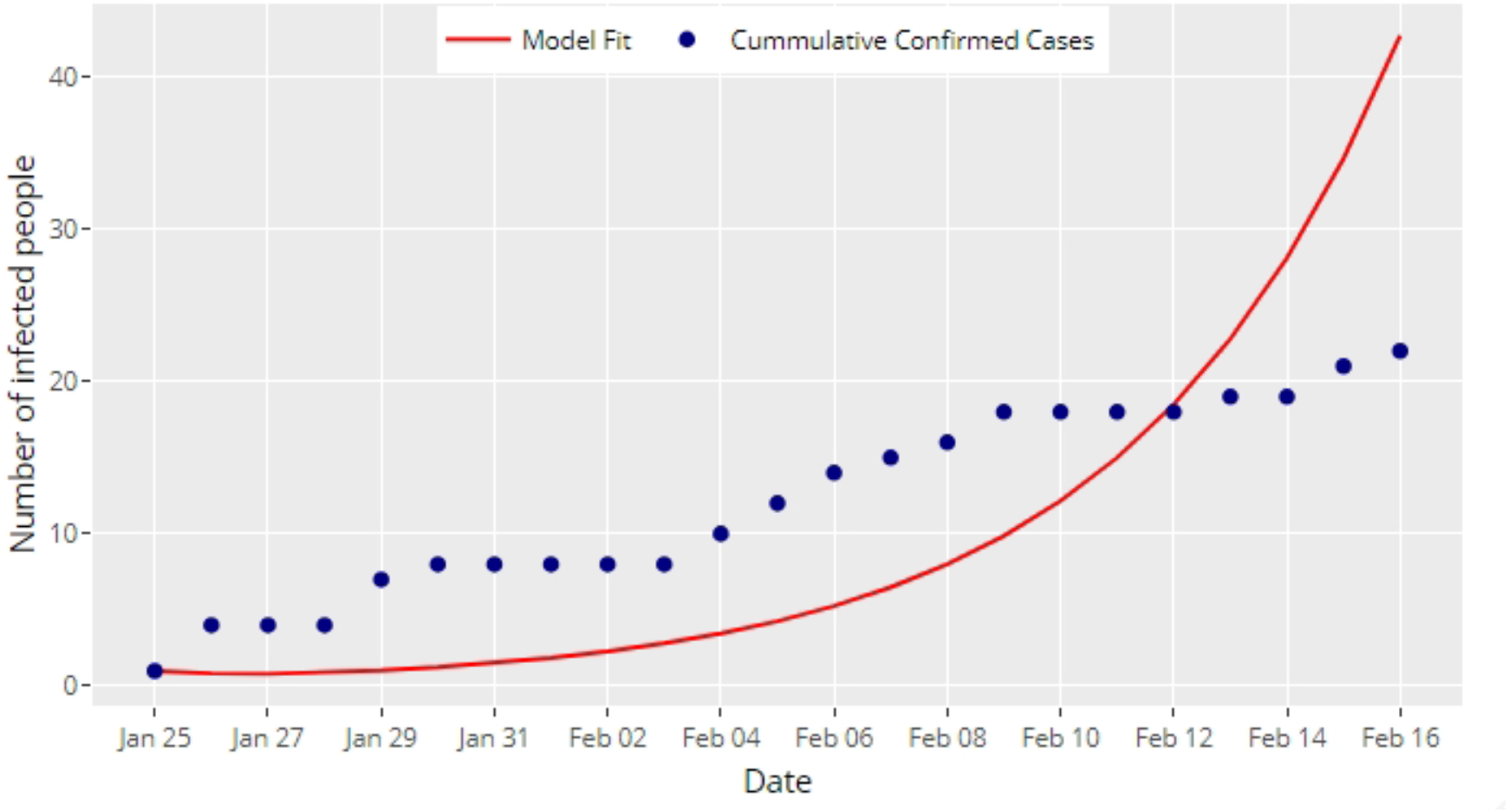
**The number of cumulative confirmed cases and model fit by date, 25 January to 16 February 2020, Malaysia**

**Figure 4:**
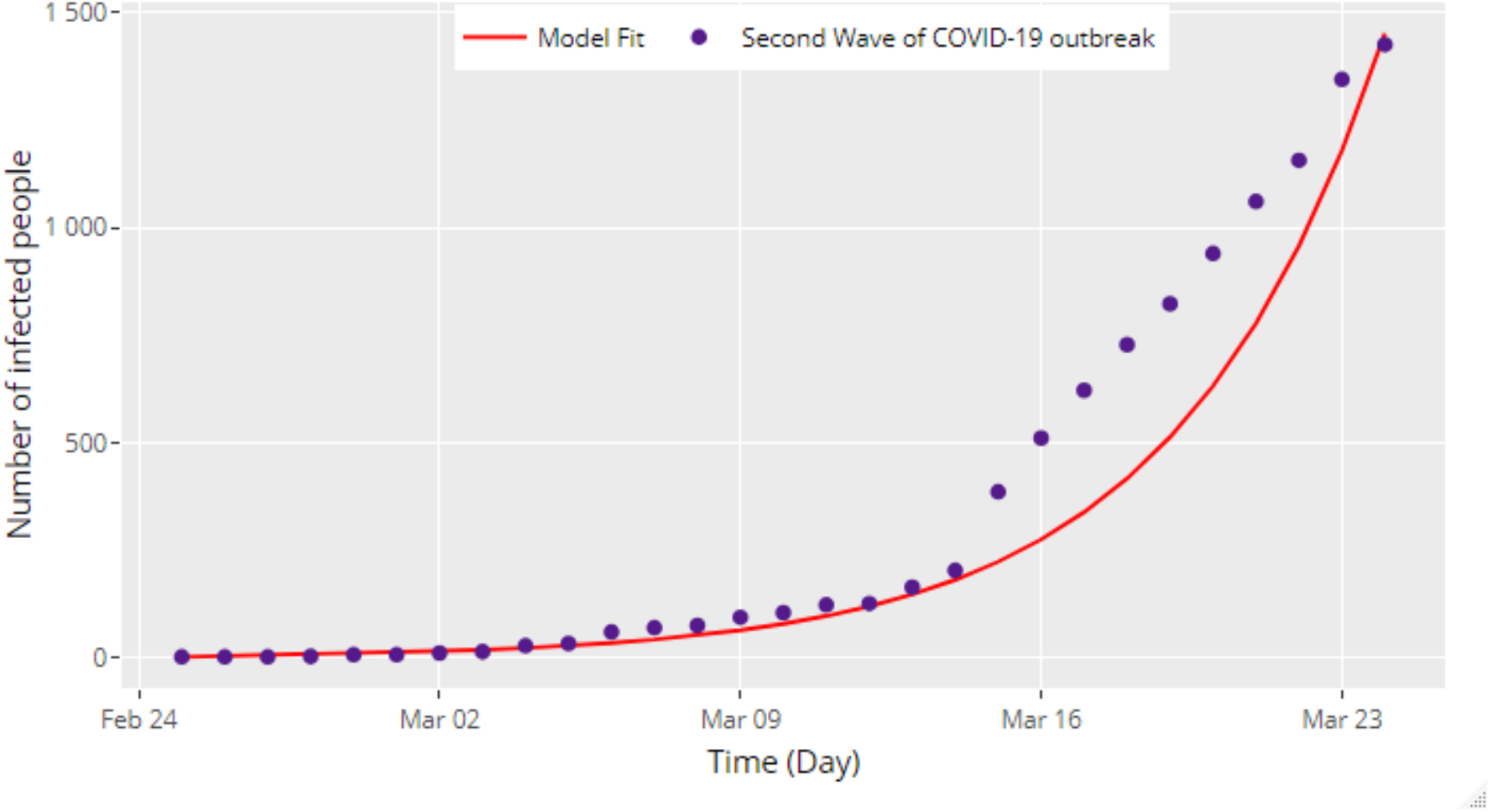
**The Second Wave of COVID-19 outbreak and model fit by date, 25 February to 18 March 2020, Malaysia**

The difference between the model fit and the cumulative confirmed cases is 19. These 19 cases are assumed as undetected cases within the period. These 19 cases used as the initial value to simulate the second wave of COVID-19 outbreak in Malaysia which is from 25 February to 18 March 2020.

## Result

The biggest transmission cluster in Malaysia is the Sri Petaling tabligh gathering. The event was held on 27^th^ February to 1^st^ March 2020 in Sri Petaling, involving a total of 16,000 participants from several countries. Among those participants, 14,500 are from Malaysia. Few days after the event, Brunei reported their first case of COVID-19 which was linked to the Sri Petaling tabligh gathering. In order to include this new information with regards to the transmission dynamics of COVID-19 in Malaysia, we simulated the disease spread locally for the Sri Petaling tabligh event where the initial values for exposed and infected populations will be identified. By using the SEIR model formulated above, the total population is assigned to 14500 which represents the total Malaysian participants. The sum of the initial exposed and infected in this group, *E*_*0*_ *+ I*_*0*_, was assumed to be equal to 40, of which is the total infected Malaysian linked to the tabligh gathering as reported on the 13^th^ March 2020. The model is simulated and compared with the actual cumulative confirmed cases from the tabligh gathering, and the root mean square error (RMSE) is calculated as an indicator to base on for the best combination of *E*_*0*_ and *I*_*0*_ Table 2 shows the RMSE for each combination.

**Table 2.**
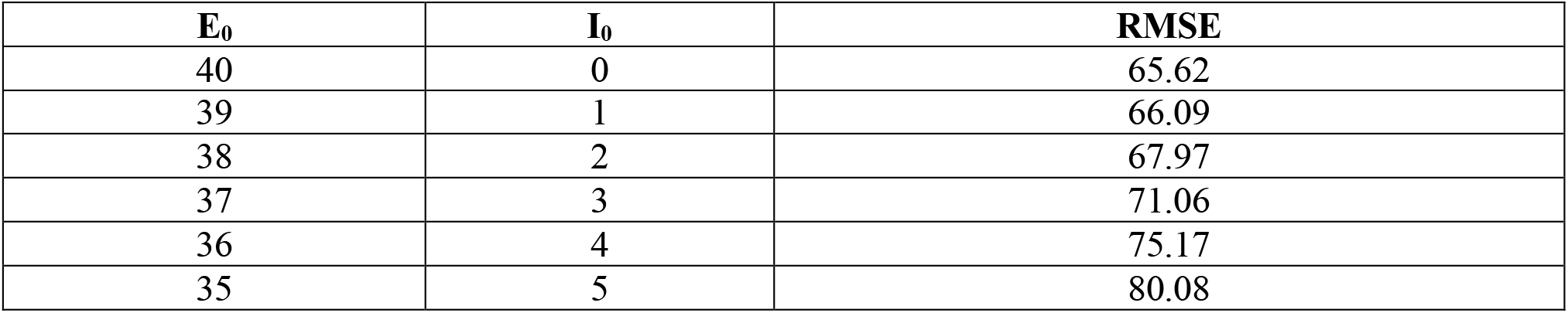
**Root Mean Square Error for the combination of E**_0_ **and I**_0_

From Table 2, the smallest RMSE value is 65.62, which is when *E*_*0*_ and *I*_*0*_ are 40 and 0 respectively. Based on this output, it gives an estimation for the event that there is a total of 40 participants who were infectious with or without symptoms. The event is assumed to be a closed population and did not take any efficient precautions of monitoring the temperature and the travel history of their participants. The model fitting of the total infected of the event is shown in figure 5.

**Figure 5:**
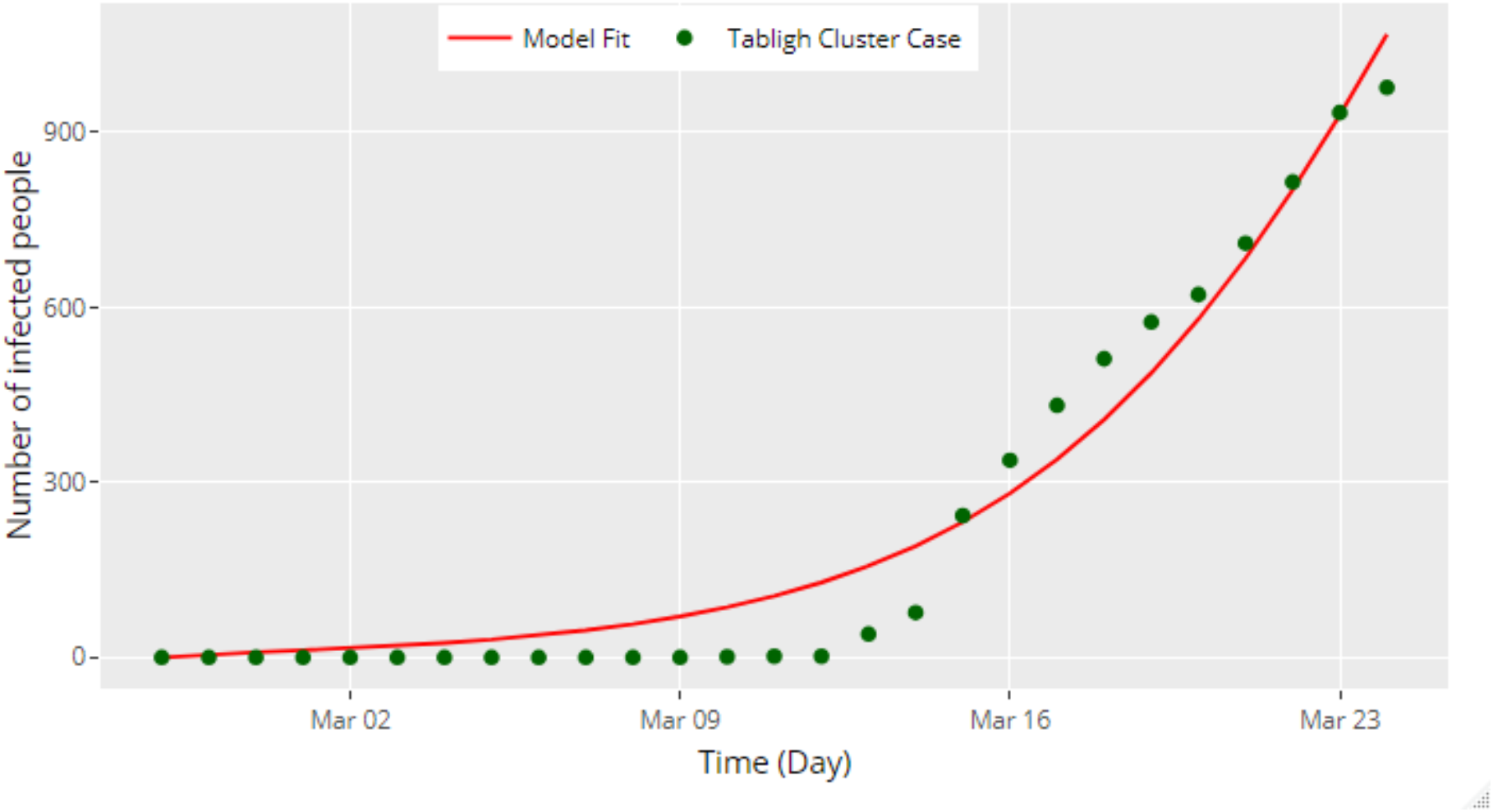
**The Sri Petaling Tabligh gathering infected and model fit by date, 27 February to 24 March 2020, Malaysia**

Figure 6 depicted the simulation of the number of infected from the end of February to May 2020. The simulation estimated more than 2000 participant will be infected and the peak will occur in early April if control is not imposed on the participants. Until 24 March, a total of 983 participants of the tabligh gathering is tested positive and 6 death cases are reported from the 983 participants. To stop the transmission chain and control the outbreak, the undetected 5% of participants need to be identified and to perform isolation. The contact tracing of those participants to also be identified and continue to quarantine at home.

**Figure 6:**
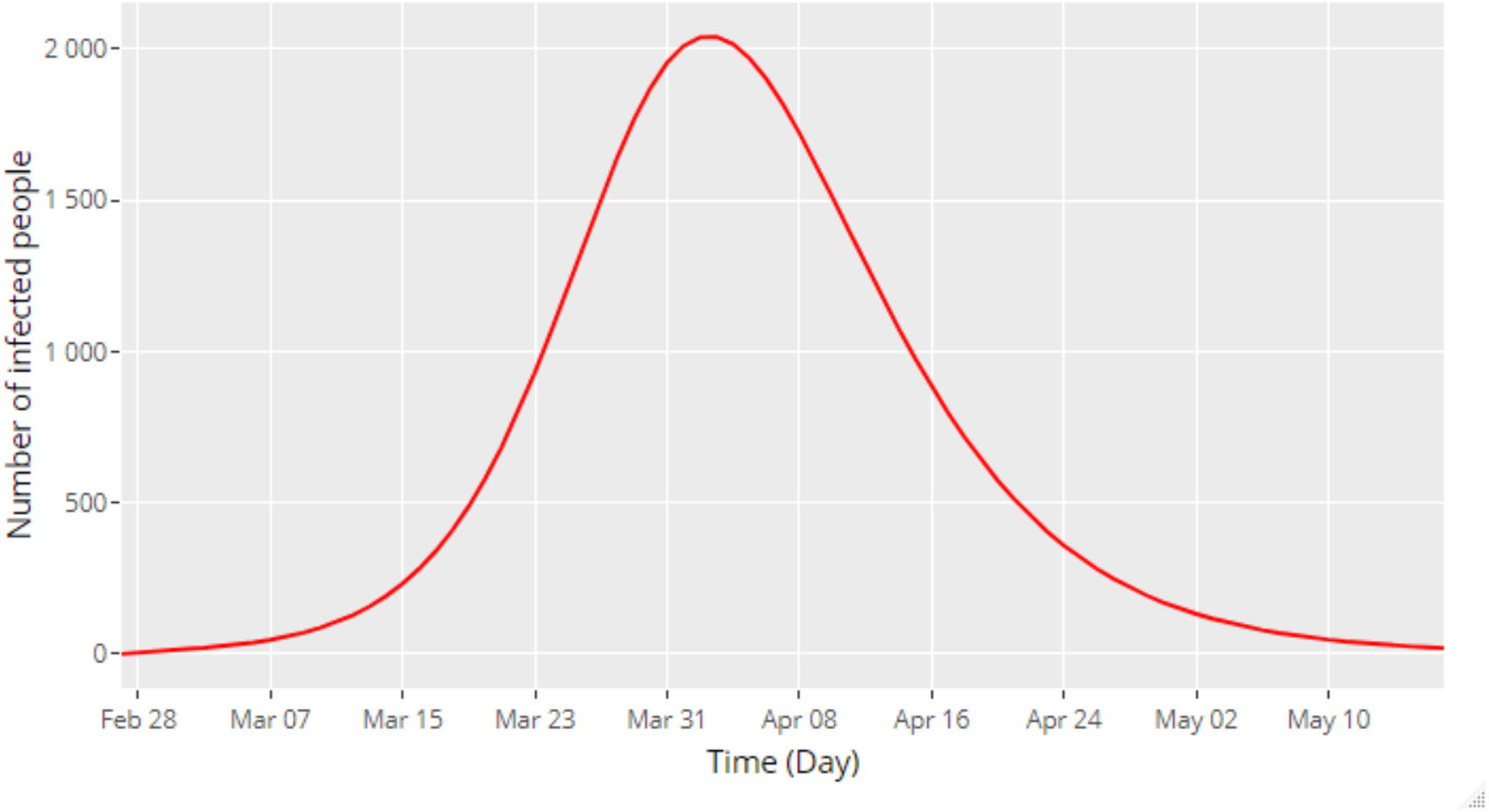
**The Simulation the total infected of Sri Petaling Tabligh Gathering**

In figure 7, the simulation is run for 150 days for Malaysia as a whole and it is observed that if there is no control measure taken then the disease will run its course from 25 February 2020 until the middle of May 2020 when the maximum number of infected cases is reached. This means that if there is no control intervention imposed on the situation then the disease will be able to infect more than 4 million in the country. The parameter value used is as listed in table 1.

**Figure 7.**
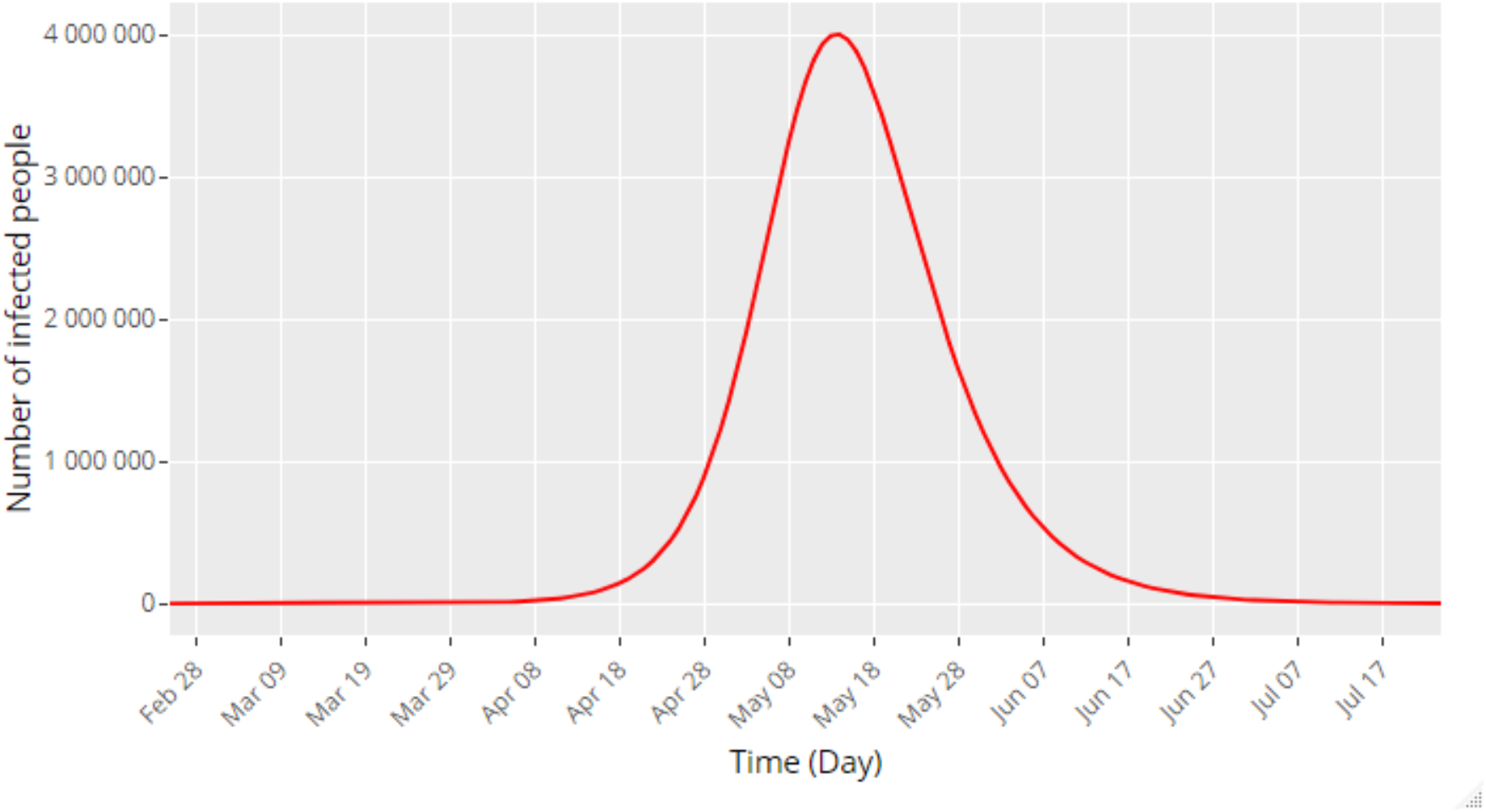
**Simulation of SEIR model on Malaysia population in 150 days**

The basic reproduction number, *R*_*0*_ is derived by using the next-generation matrix approach, the equation is shown in equation 5

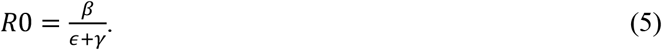

Using equation 5, the *R*_*0*_ is calculated as 3.74. which is acceptable if we were to compare with the studies by [11] who gives a range of the basic reproduction number for COVID-19 to be in the range from 1.4 to 6.49. Malaysia currently under the acceleration stage since the number of cases reported increases rapidly, hence the *R*_*0*_ is higher at this stage. To eradicate the disease among the community, the *R*_*0*_ needs to be less than 1. Taking efficient control measures and precaution such as social distancing, movement restriction, it can reduce the *R*_*0*_ since the contact rate between susceptible and infected will be decreased when such control is in place.

## Discussion

The Prime Minister of Malaysia has promulgated the Movement Control (MCO) order on 16 March 2020 and this order is effective from 18 March to 31 March, a total of 14 days [12]. This order aims to reduce the contact between people by forbidding several types of activities such as sport, social, cultural and religious. Only the business that provides basic daily needs is allowed to operate within this period. As mentioned before, *R*_*0*_ can be reduced by decreasing the contact rate between people, hence the transmission rate will be decreased. This is because the transmission is affected by the contact rate between susceptible and infected based on the assumption of mass action law [13].

## Conclusion

The COVID-19 SEIR model was formulated and simulated using the parameters values obtained from trusted sources. The model is found to be accurate as it is able to predict the second confirmed case in Malaysia as reported on February 7 by the MoH [5]. One of the parameters used, namely the incubation period, was actually estimated by [2] based on 88 confirmed cases detected outside of Wuhan, China. Our simulation has managed to verify the proposed incubation period of 6.5 days at which according to the MoH report [5], the second case has developed symptoms on the 1st February 2020 which is in agreement with the prediction of our model. With the same model, we have simulated the disease transmission amongst the Malaysian participants of the Sri Petaling tabligh event. We have seen how fast the disease spreads and most likely will affect the Malaysia total population. We have also shown through the simulation the seriousness of the spread of the disease if there is no control measure is taken.

## Data Availability

The data obtained are publicly available from trusted sources

Workflow of Next Generation Matrix Approach (NGM)

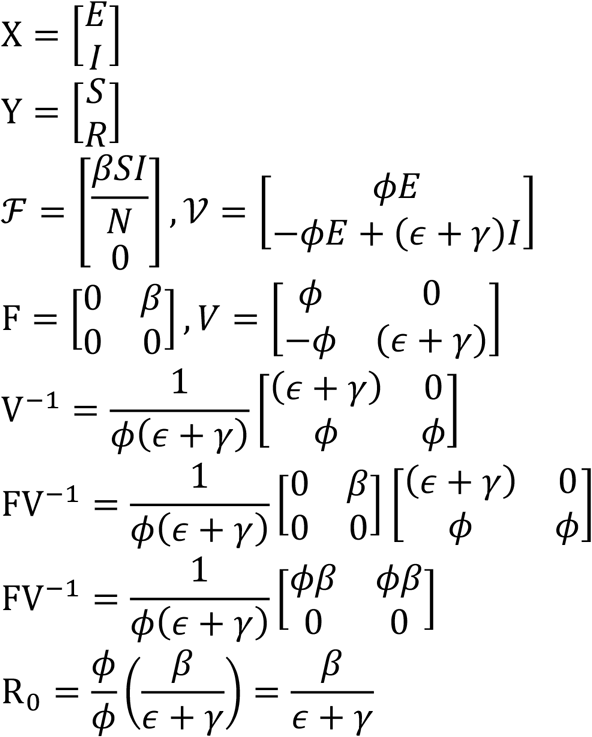

